# Preliminary study to identify severe from moderate cases of COVID-19 using NLR&RDW-SD combination parameter

**DOI:** 10.1101/2020.04.09.20058594

**Authors:** Changzheng Wang, Rongrong Deng, Liyao Gou, Zhongxiao Fu, Xiaomei Zhang, Feng Shao, Guanzhen Wang, Weiyang Fu, Jianping Xiao, Xiao Ding, Tao Li, Xiulin Xiao, Chengbin Li

## Abstract

**Objectives:** Investigate the characteristics and rules of hematology changes in patients with COVID-19, and explore the possibility to identify moderate and severe patients using conventional hematology parameters or combined parameters.

**Methods:** The clinical data of 45 moderate and severe type patients with SARS-CoV-2 infections in Jingzhou Central Hospital from January 23 to February 13, 2020 were collected. The epidemiological indexes, clinical symptoms and laboratory test results of the patients were retrospectively analyzed. Those parameters with significant differences between the two groups were analyzed, and the combination parameters with best diagnostic performance were selected using the LDA method.

**Results:** Of the 45 patients with COVID-19 (35 moderate and 10 severe cases), 23 were male and 22 female, aged 16-62 years. The most common clinical symptoms were fever (89%) and dry cough (60%). As the disease progressed, WBC, Neu#, NLR, PLR, RDW-CV and RDW-SD parameters in the severe group were significantly higher than that in the moderate group (*P*<0.05); meanwhile, Lym#, Eos#, HFC%, RBC, HGB and HCT parameters in the severe group were significantly lower than that in the moderate group (*P*<0.05). For NLR, the AUC, the best cut-off value, the sensitivity and the specificity were 0.890, 13.39, 83.3% and 82.4% respectively, and for PLR, the AUC, the best cut-off, the sensitivity and the specificity were 0.842, 267.03, 83.3% and 74.0% respectively. The combined parameter NLR&RDW-SD had the best diagnostic efficiency (AUC was 0.938) and when the cut-off value was 1.046, the sensitivity and the specificity were 90.0% and 84.7% respectively, followed by the fitting parameter NLR&RDW-CV (AUC = 0.923). When the cut-off value was 0.62, the sensitivity and the specificity for distinguishing severe type from moderate cases of COVID-19 were 90.0% and 82.4% respectively.

**Conclusions:** The combined parameter NLR&RDW-SD is the best hematology index and can help clinicians to predict the severity of COVID-19 patients, and it can be used as a useful indicator to help prevent and control the epidemic.

## Introduction

Coronavirus was first isolated and named in the 1960s. It is a zoonotic RNA virus that can spread between animals and humans. It can cause respiratory and intestinal infections in mammals and birds. There are currently seven known coronaviruses that can infect humans, four of which are common pathogens of human colds, which usually do not cause serious illness. Patients show common cold symptoms such as fever and swelling of the throat [1]. Coronavirus initially got really attention because of severe acute respiratory syndrome (SARS) caused by the SARS coronavirus (SARS-CoV) that broke out in Asia in 2002-2003, with more than 8,000 people infected and a mortality rate of approximately 9.6% [4-6, 20], and subsequently the Middle East respiratory syndrome (MERS) caused by the Middle East respiratory syndrome coronavirus (MERS-CoV) that broke out again in 2012 in the Middle East, Africa and other regions, with more than 2,000 diagnosed cases and a mortality rate of about 34.4% [7, 21]. The third fatal coronavirus is novel coronavirus (SARS-CoV-2) that cause novel coronavirus pneumonia (COVID-19) which first broke out in Wuhan, China in December 2019 [1, 8]. Fever, dry cough and fatigue are the main manifestations. Severe patients often have dyspnea and/or hypoxemia one week after the onset of symptoms. In severe cases, they can quickly progress to acute respiratory distress syndrome (ARDS), septic shock, metabolic acidosis difficult to correct, coagulation dysfunction, and multiple organ failure [9-11]. As of March 16, 2020, there had been more than 80,000 confirmed cases in various provinces and cities in China, with a mortality rate of approximately 3.97%. The number of confirmed patients outside China has increased rapidly in the world. About 24,000 were diagnosed on March 8, but by March 16, more than 86,000 cases had been diagnosed. The mortality rate also increased from 1.96% to 3.92% [19].

According to the Guidelines for the Diagnosis and Treatment of Novel Coronavirus (2019-nCoV) Infection (trial version 6) [2] (hereinafter referred to as “Diagnosis and Treatment Guidelines”) issued by the National Health Commission of the People’s Republic of China, COVID-19 is divided into four types, mild, moderate, severe and critical based on the clinical manifestations of the patient’s disease and treated with different measures. Patients with mild clinical manifestations may not initially need to be admitted for treatment, but may show respiratory symptoms within the second week, so all patients should be closely monitored. WHO reports that about 80% of infected people are mild to moderate infected (including those with or without pneumonia), 13.8% of infected people have severe infections, and 6.1% of infected people have critical illness [14]. A meta-analysis of more than 50,000 cases showed that severe cases accounted for 18.1% in all infected patients [15]. It is also reported [13] that patients with mild to moderate infection, severe infection and critical infection accounted for 80.9%, 13.8% and 4.7% in confirmed cases respectively. Experts from China had reported that approximately 26.1% -32.0% of confirmed cases would develop into severe or critical cases [9, 11], and the fatality rate of critical cases would reach an alarming level of 61.5% [12]. It is reported that with the increase of the age of infected patients, the mortality rate goes up, and the crude mortality rate in people over 80 years old is 21.9% [14], so the identification and diagnosis of severe or critical patients is very important. Routine hematology tests are the most basically and most commonly carried out in laboratories. Patients usually have to take a test every day or every two days. If it can play an important role in severe cases identification, it will provide clinicians with more auxiliary diagnostic information.

The Diagnosis and Treatment Guidelines (trial version 6) [2] clearly pointed out that the total number of peripheral white blood cells was normal or decreased and the lymphocyte count was reduced in the early stage of onset. Li et al [16] conducted a descriptive and predictive study and found that lymphocyte percentage (Lym%) was inversely related to the severity and prognosis of patients, which could be used to predict the severity and prognosis of patients with COVID-19. It indicates that some changes in the peripheral blood will occur in patients with SARS-CoV-2 infections. These changes have the potentiality to provide clues or guidance for the diagnosis, treatment and prognosis for COVID-19 patients. BC-6900 is newest hematology analyzer of MINDRAY Medical International Co., Ltd (Shenzhen, China). It uses the principle of nucleic acid fluorescence staining and flow cytometry to detect white blood cells, red blood cells and platelets in peripheral venous blood in three dimensions. The blood cells are identified and quantitatively analyzed according to the volume of the cells and the complexity of the contents, as well as the nucleic acid content. In addition to providing the most routine hematology parameters, it can also quantitatively detect immature granulocytes, nucleated red blood cells and naive platelets, as well as blast cells. Based on the analysis of the results from BC-6900, this study intend to discover the characteristic changes of the peripheral blood and explore the value of hematology routine parameters in the diagnosis and treatment for COVID-19 patients.

This study focuses on the identification of critical COVID-19 patients using hematology routine parameters. In order to explore the value of hematology routine parameters, we retrospectively analyzed the epidemiological and laboratory test results of 45 moderate and severe cases. The differences between the different groups found through the most routine laboratory tests were analyzed in order to provide valuable help for clinicians to diagnose and treat this disease more effectively.

## Material and Methods

### Patients

In this study, all data of 45 hospitalized cases were collected between Jan 23, 2020 and Feb 13, 2020 from the Department of Laboratory Medicine, Jingzhou central Hospital, The 2^nd^ Clinical Medical College, Yangtze University (Jingzhou, Hubei province, People’s Republic of China). Among all 45 patients, 35 cases were moderate and 10 cases were severe type. All patients were confirmed by viral detections using novel coronavirus 2019-nCoV nucleic acid detection kit (fluorescent PCR method, Shanghai BioGerm Medical Biotechnology Co., Ltd). And all patients were ruled out of coinfections by other respiratory virus including respiratory syncytial virus, adenovirus, influenza virus A, influenza virus B, parainfluenza virus, chlamydia pneumoniae, legionella pneumoniae and mycoplasma pneumoniae using serological method. All case were diagnosed and classified according to the Diagnosis and Treatment Guidelines (Trial Sixth Edition). The clinical standards for the identification of moderate and severe patients are as follows: 1) moderate, with fever, respiratory track symptoms and pneumonia imaging; 2) severe, having any of the following conditions beside the symptoms and sign of moderate: a) respiratory distress, RR ≥30 times/minute; b) oxygen saturation ≤93% under rest state; c) arterial blood oxygen partial pressure (PaO2)/oxygen concentration (FiO2) ≤300mmHg and d) lung imaging progress >50% in the short term (24∼48hours).

### Data collection

The epidemiological characteristics information (including recent exposure history, such as the travel history and contacting with patients with fever or respiratory symptoms from other cities in Hubei province or confirmed cases within two weeks) and the basic information such as gender, age, clinical symptoms and signs were collected from the admission records. All the laboratory data including complete blood count (BC-6900, Mindray), serum biochemistry and coagulation test were collected from the laboratory information system (LIS).

In this study, total 161 results detected on the Mindray BC-6900 hematology analyzer and of biochemistry as well as coagulation tests were collected from 45 patients with SARS-CoV-2 infection between Jan 23, 2020 and Feb 13, 2020. All the laboratory test results were divided into two groups based on the sources of the samples, from moderate or severe patients, on which retrospective analysis and comparative analysis were performed.

### Statistical analysis

In this study, SPSS statistics software (*version* 19.0) was used for data statistics and mapping. Age was represented in median (range), and others demographics and clinical characteristics were expressed in frequency and percentage. The significance was tested by chi square or Fisher’s exact test. The quantized variables of blood parameters were expressed as mean ± standard deviation. The significance between the two groups was tested by student’s t-test. *P*<0.05 was considered statistically significant in all statistical analyses. Linear discriminant analysis (LDA) was employed to perform linear combination of each two parameters and extract the best data features to distinguish moderate and severe cases of COVID-19 patients. The diagnostic values of valuable parameters for differential mild and severe cases of COVID-19 patients were assessed by receiver operating characteristic (ROC) and area under the ROC curve (AUC).

### LDA for combined parameters

The combined parameters were analyzed using LDA. LDA is a supervised learning model, also known as Fisher’s linear discriminant (FLD) [27]. The principle of LDA is: by multi-parameter linear combination, the high-dimensional pattern samples composed of multi-parameters are projected to the optimal discrimination vector space to achieve the effect of extracting classification information and compressing the feature space dimension. The new subspace has the largest inter-class distance and the smallest intra-class distance, that is, the pattern has the best separability in this space. Therefore, it is an effective feature extraction method.

## Results

### Demographic and clinical characteristics of COVID-19 patients

Forty-five confirmed COVID-19 patients were divided into moderate (35 cases, 77.8%) and severe (10 cases, 22.2%) infection groups (Table 1). There was no significant difference in median age between the two groups (*P*> 0.05). Of the patients, 23 were males (51.1%) and 22 females (48.9%). There was also no significant difference in gender composition between the two groups of patients (*P*> 0.05). In all cases, there were total 26 patients who had been to Wuhan within 2 weeks before their hospitalization, and of theses 26 patients 20 were moderate patients (57.1%) and 6 severe patients (60%). There was no significant difference in Wuhan contacting history between the two groups (*P*> 0.05). Only 3 (6.7%) patients had ever visited the South China Seafood Market, and all these three patients were moderate patients. It is worth noting that there were 4 patients with hypertension-based diseases, of which 3 (30%) were severe patients. Of all the 45 patients, 40 (89%), 27 (60%), 19 (42%), 15 (33%) and 13(28.9%) cases had fever, dry cough, fatigue, chills and myalgia respectively, and there was no significant difference between the two groups (*P*> 0.05).

**Table 1.**
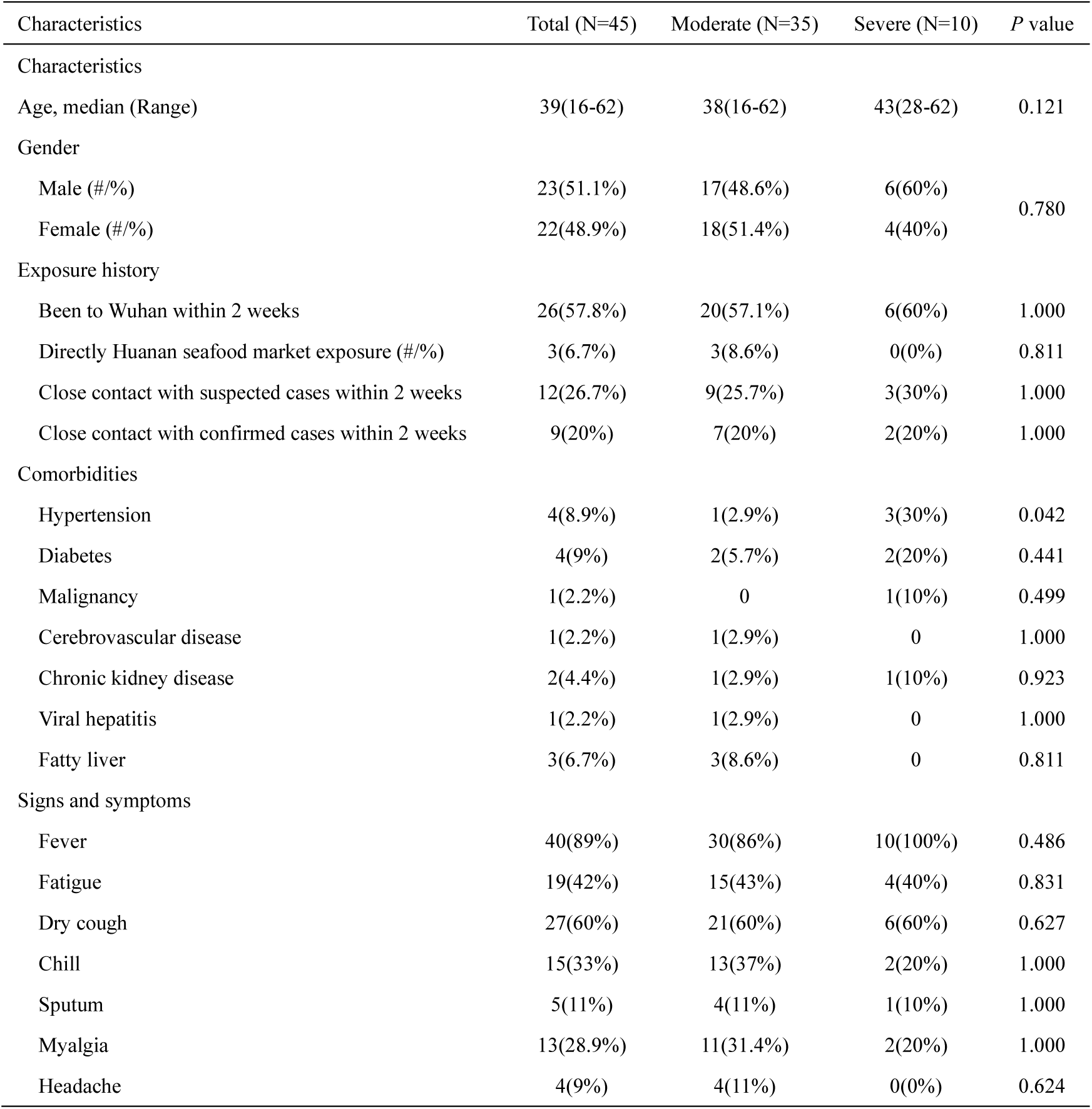
Demographics and clinical characteristics of COVID-19 patients.

| Characteristics | Total (N=45) | Moderate (N=35) | Severe (N=10) | P value |
| --- | --- | --- | --- | --- |
| Characteristics |  |  |  |  |
| Age, median (Range) | 39(16-62) | 38(16-62) | 43(28-62) | 0.121 |
| Gender |  |  |  |  |
| Male (#/%) | 23(51.1%) | 17(48.6%) | 6(60%) | 0.780 |
| Female (#/%) | 22(48.9%) | 18(51.4%) | 4(40%) |  |
| Exposure history |  |  |  |  |
| Been to Wuhan within 2 weeks | 26(57.8%) | 20(57.1%) | 6(60%) | 1.000 |
| Directly Huanan seafood market exposure (#/%) | 3(6.7%) | 3(8.6%) | 0(0%) | 0.811 |
| Close contact with suspected cases within 2 weeks | 12(26.7%) | 9(25.7%) | 3(30%) | 1.000 |
| Close contact with confirmed cases within 2 weeks | 9(20%) | 7(20%) | 2(20%) | 1.000 |
| Comorbidities |  |  |  |  |
| Hypertension | 4(8.9%) | 1(2.9%) | 3(30%) | 0.042 |
| Diabetes | 4(9%) | 2(5.7%) | 2(20%) | 0.441 |
| Malignancy | 1(2.2%) | 0 | 1(10%) | 0.499 |
| Cerebrovascular disease | 1(2.2%) | 1(2.9%) | 0 | 1.000 |
| Chronic kidney disease | 2(4.4%) | 1(2.9%) | 1(10%) | 0.923 |
| Viral hepatitis | 1(2.2%) | 1(2.9%) | 0 | 1.000 |
| Fatty liver | 3(6.7%) | 3(8.6%) | 0 | 0.811 |
| Signs and symptoms |  |  |  |  |
| Fever | 40(89%) | 30(86%) | 10(100%) | 0.486 |
| Fatigue | 19(42%) | 15(43%) | 4(40%) | 0.831 |
| Dry cough | 27(60%) | 21(60%) | 6(60%) | 0.627 |
| Chill | 15(33%) | 13(37%) | 2(20%) | 1.000 |
| Sputum | 5(11%) | 4(11%) | 1(10%) | 1.000 |
| Myalgia | 13(28.9%) | 11(31.4%) | 2(20%) | 1.000 |
| Headache | 4(9%) | 4(11%) | 0(0%) | 0.624 |

### Hematology Findings of COVID-19 patients

Total 161 venous blood samples anticoagulated by EDTA-K2 were collected from those 45 patients between Jan 23, 2020 and Feb 13, 2020 in our laboratory. Among them, 131 venous blood samples were collected from 35 moderate patients, and the other 30 from 10 severe patients. The hematology characteristics of those samples from the two groups were presented in Table 2. As the disease progressed, WBC, Neu#, NLR, PLR, RDW-CV and RDW-SD parameters in the severe group were significantly higher than that in the moderate group (*P*<0.05); meanwhile, Lym#, Eos#, HFC%, RBC, HGB and HCT parameters in the severe group were significantly lower than that in the moderate group (*P*<0.05). The box-plots for those significant parameters were all presented in Figure 1.

**Table 2.** Hematology findings of venous samples taken from COVID-19 patients.

| Parameters | Total (N=161) | Moderate (N=131) | Severe (N=30) | Levene test* (P) | t-test(P) |
| --- | --- | --- | --- | --- | --- |
| WBC, $\times 10^9/L$ | 9.12±4.41 | 8.59±4.01 | 11.46±5.32 | 0.065 | 0.001 |
| Neu#, $\times 10^9/L$ | 7.41±4.34 | 6.69±3.83 | 10.51±5.10 | 0.051 | 0.000 |
| Lym#, $\times 10^9/L$ | 1.18±0.79 | 1.34±0.78 | 0.50±0.39 | 0.000 | 0.000 |
| Mon#, $\times 10^9/L$ | 0.49±0.25 | 0.50±0.25 | 0.43±0.27 | 0.544 | 0.205 |
| Eos#, $\times 10^9/L$ | 0.04±0.06 | 0.04±0.06 | 0.00±0.01 | 0.000 | 0.000 |
| Bas#, $\times 10^9/L$ | 0.02±0.01 | 0.02±0.01 | 0.02±0.02 | 0.004 | 0.027 |
| IMG%, % | 1.42±1.85 | 1.45±1.97 | 1.30±1.23 | 0.213 | 0.691 |
| HFC%, % | 0.49±0.49 | 0.52±0.53 | 0.36±0.27 | 0.023 | 0.026 |
| NLR, % | 12.02±13.94 | 7.93±8.36 | 29.9±18.7 | 0.000 | 0.000 |
| PLR, % | 316.46±309.28 | 238.8±196.0 | 655.6±457.4 | 0.000 | 0.000 |
| RBC, $\times 10^{12}/L$ | 4.28±0.58 | 4.36±0.46 | 3.89±0.88 | 0.000 | 0.007 |
| HGB, g/L | 132.22±16.64 | 134.5±12.1 | 122.3±27.3 | 0.000 | 0.023 |
| HCT, % | 39.36±4.86 | 40.1±3.54 | 36.3±7.88 | 0.000 | 0.015 |
| MCV, fl | 92.11±3.33 | 91.88±3.43 | 93.11±2.64 | 0.077 | 0.034 |
| MCH, pg | 30.95±1.28 | 30.85±1.35 | 31.37±0.82 | 0.009 | 0.008 |
| MCHC, g/L | 335.99±6.61 | 335.8±6.88 | 337.0±5.25 | 0.064 | 0.354 |
| RDW-CV, % | 12.34±0.51 | 12.29±0.46 | 12.59±0.65 | 0.001 | 0.020 |
| RDW-SD | 39.84±1.73 | 39.52±1.54 | 41.21±1.84 | 0.104 | 0.000 |
| PLT, $\times 10^9/L$ | 220.42±70.82 | 222.7±73.01 | 210.6±60.43 | 0.387 | 0.403 |
| MPV, fl | 9.83±1.19 | 9.74±1.22 | 10.17±0.98 | 0.168 | 0.075 |
| PDW, % | 16.25±0.31 | 16.23±0.31 | 16.36±0.30 | 0.487 | 0.040 |
| PCT, % | 0.21±0.07 | 0.21±0.07 | 0.21±0.05 | 0.112 | 0.760 |
| P-LCR, % | 25.04±7.9 | 24.55±8.11 | 27.20±6.54 | 0.166 | 0.097 |

\*: Levene test was used for the homogeneity of variance test. White blood cell count (WBC), Neutrophil count (Neu#), Lymphocyte count (Lym#), Monocyte count (Mon#), Eosinophil count (Eos#), Basophil count (Bas#), Immature Granulocyte Percentage (IMG%), High Fluorescent Cell Percentage(HFC%), Neutrophil-to-Lymphocyte Ratio(NLR), Platelet-to-Lymphocyte Ratio (PLR), Red blood cell count (RBC), Hemoglobin (HGB), Hematocrit (HCT), Mean corpuscular volume (MCV), Mean Corpuscular Hemoglobin (MCH), Mean Corpuscular Hemoglobin Concentration (MCHC), Red Cell volume Distribution Width-coefficient of variation (RDW-CV), Red Cell volume Distribution Width-standard deviation (RDW-SD), Platelet count (PLT), Mean platelet volume (MPV), Platelet distribution width (PDW), Platelet hematocrit (PCT), Platelet-larger cell ratio(P-LCR).

**Figure 1.**
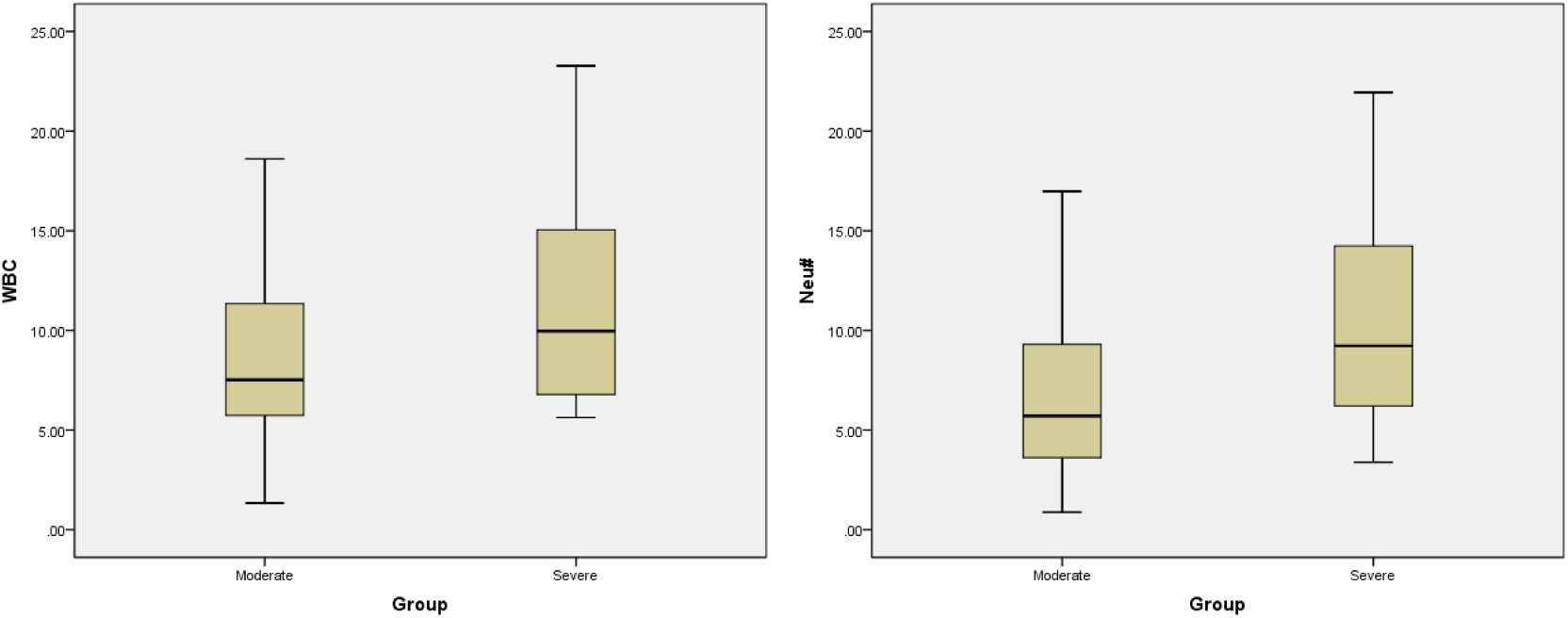
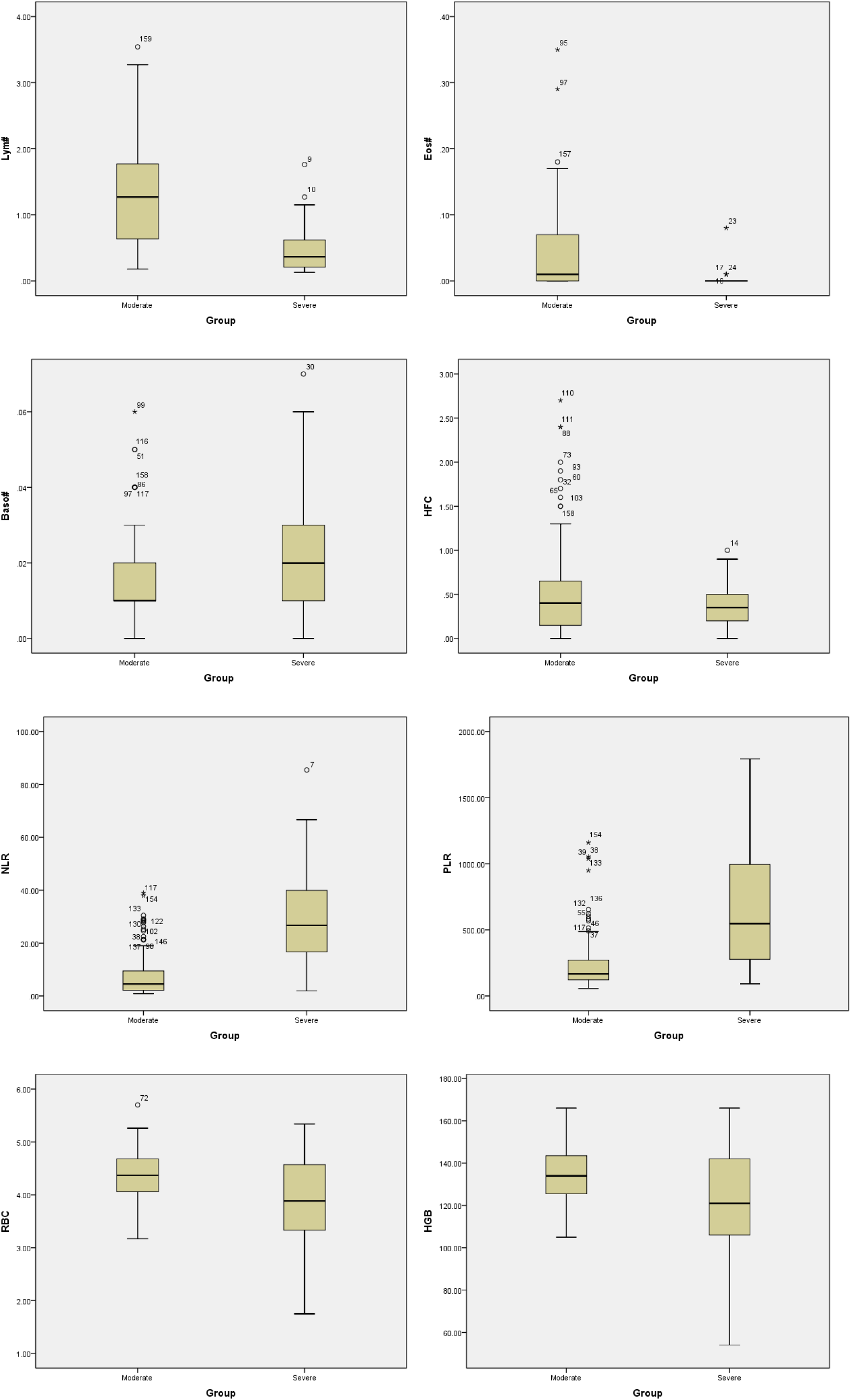
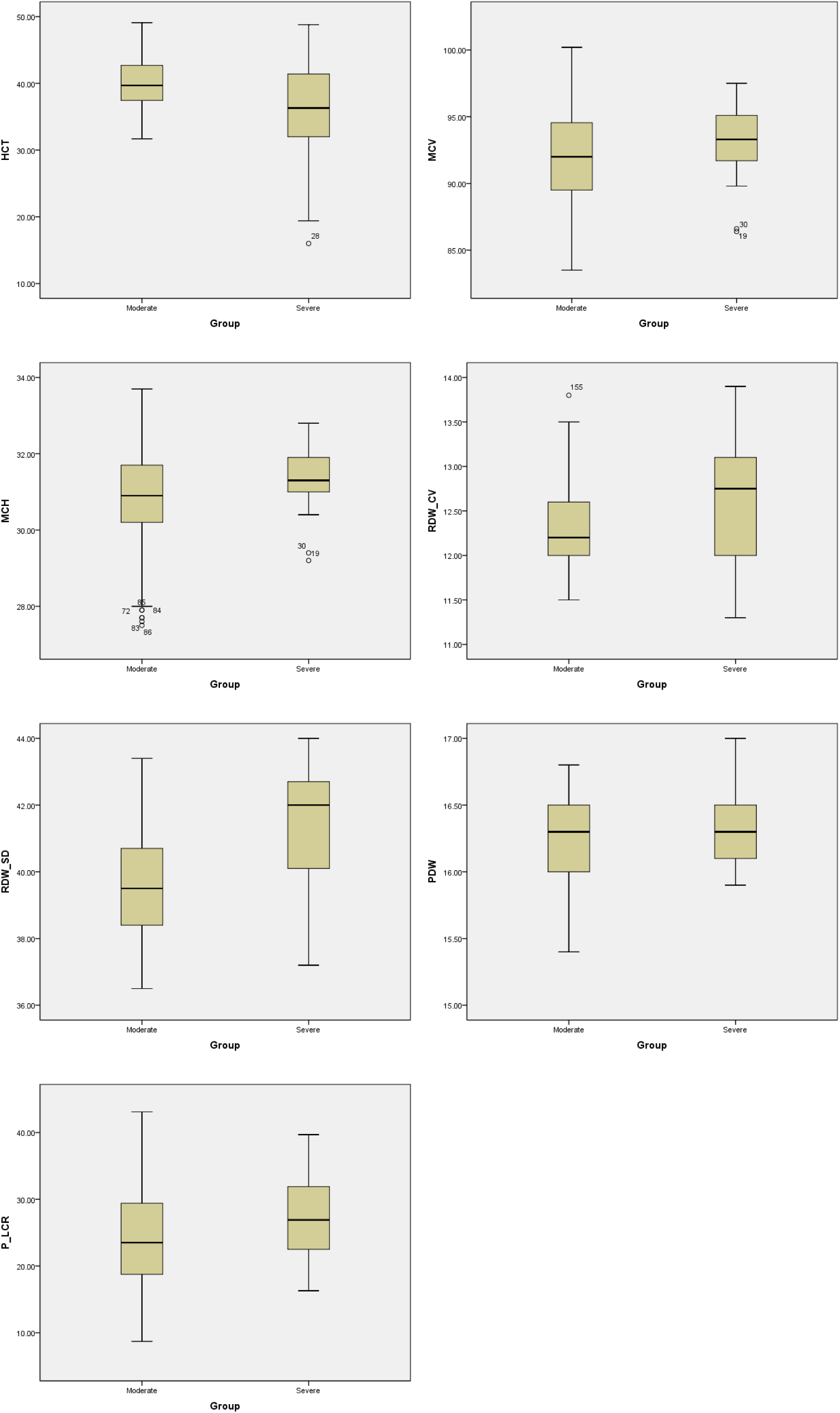
The comparison for significant CBC parameters between moderate and severe type of COVID-19 patients. The box-plots were provided and the student’s t-test was employed to compare the differences in CBC parameters between the moderate and the severe cases groups of COVID-19. \*\*\**p*<0.001.

**Figure 2.**
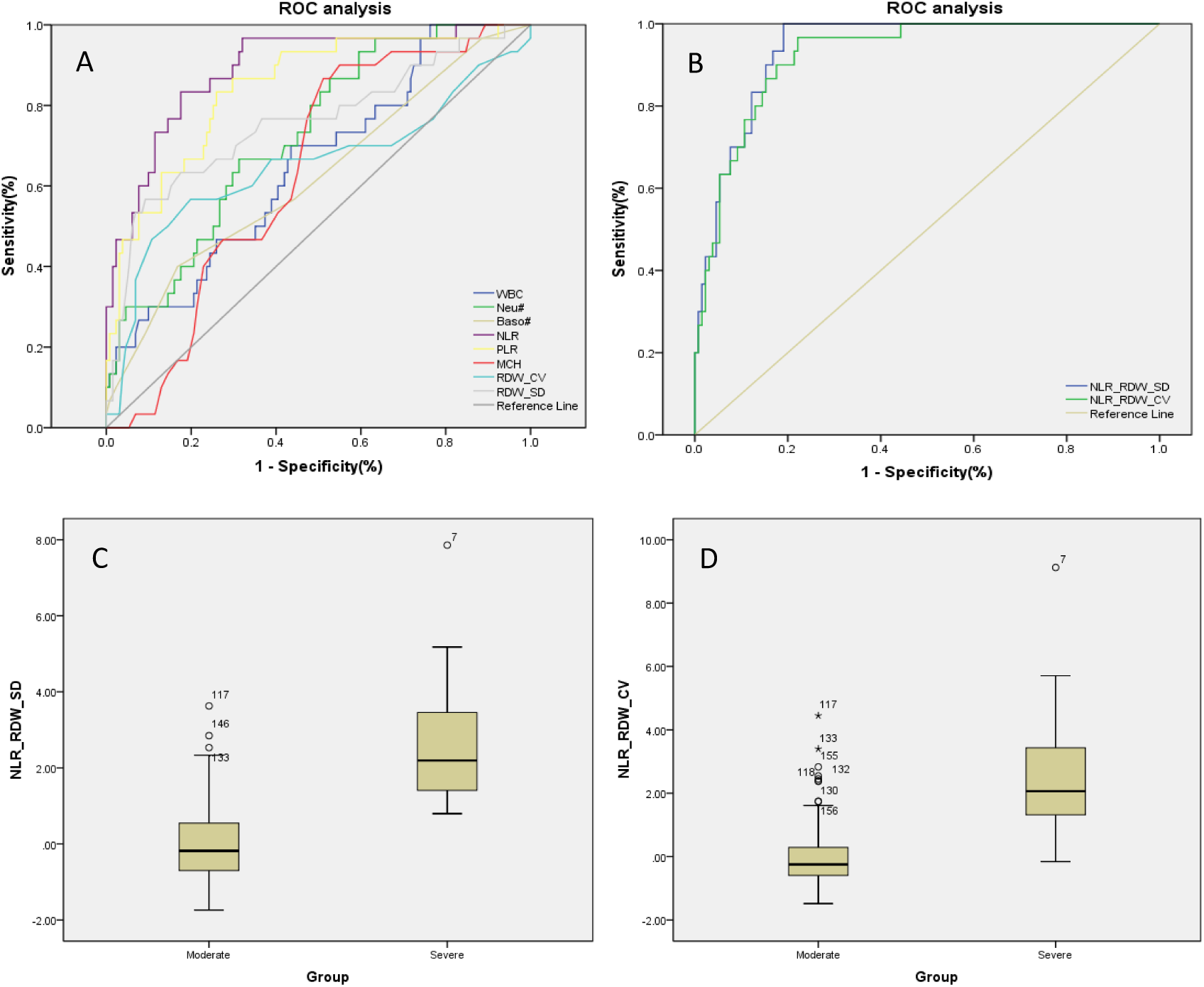
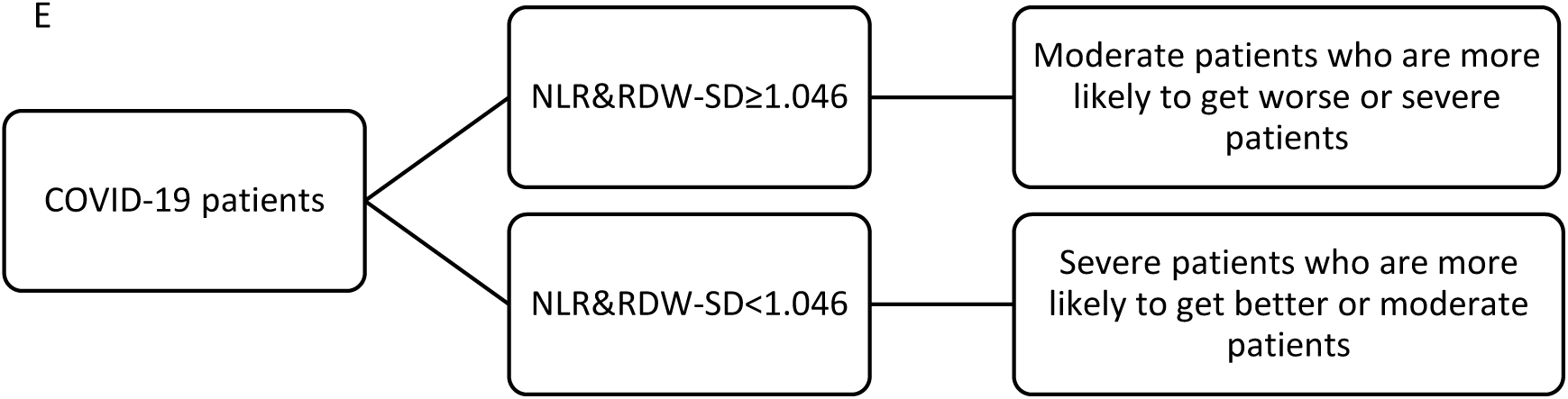
ROC analysis using single and combined parameters in the diagnosis of severe cases of COVID-19. Differentiated diagnosis of moderate and severe COVID-19 patients using different parameters. The positive sample is the blood routine result of the severe patient, and the negative sample is the blood routine result of the moderate patient. Figure A is a ROC plot that uses single parameter to identify severe from moderate patients. Figure B is a ROC plot that uses the combined parameters NLR&RDW-SD and NLR&RDW-CV to identify patients; Figures C and D are box-plots that use the combined parameters for comparison between two groups; Figure E is a recommendation management strategy for COVID-19 patients.

### Biochemical and coagulation test findings of COVID-19 patients

The Biochemical and coagulation test results of those 45 patients between Jan 23, 2020 and Feb 13, 2020 in our laboratory were presented in Table 3. The biochemical and coagulation test samples were usually collected and tested once every 1-3 days. As the disease progressed, DBil, GLO, BUN, Cr, Cys C, CK, Mb, LDH and FBG in the severe group were significantly higher than that in the moderate group (*P*<0.05); meanwhile, ALB, Na^+^ and Ca^2+^ in the severe group were significantly lower than that in the moderate group (*P*<0.05). There was no significant difference for all the coagulation test results between the two groups (*P*>0.05).

**Table 3.** Biochemical and coagulation test results of COVID-19 patients.

| Parameters | Total |  | Moderate |  | Severe |  | Levene test*<br>(P) | t-test (P) |
| --- | --- | --- | --- | --- | --- | --- | --- | --- |
| | N | Mean±SD | N | Mean±SD | N | Mean±SD | | |
| DBil, umol/L | 190 | 4.52±2.23 | 160 | 4.3±2.05 | 30 | 5.69±2.79 | 0.005 | 0.013 |
| IBil, umol/L | 190 | 8.36±3.85 | 160 | 8.31±3.85 | 30 | 8.62±3.91 | 0.576 | 0.693 |
| TBil, umol/L | 192 | 13.08±6.36 | 162 | 12.56±5.39 | 30 | 15.84±9.78 | 0.006 | 0.083 |
| TP, g/L | 196 | 66.65±8.09 | 165 | 66.66±7.42 | 31 | 66.59±11.14 | 0.024 | 0.971 |
| ALB, g/L | 202 | 35.68±5.92 | 170 | 36.29±5.68 | 32 | 32.48±6.22 | 0.865 | 0.001 |
| GLO, g/L | 196 | 30.83±5.76 | 165 | 30.21±4.36 | 31 | 34.12±9.9 | 0.000 | 0.038 |
| A/G | 196 | 1.3±1.6 | 165 | 1.23±0.23 | 31 | 1.71±4.02 | 0.000 | 0.510 |
| ALT, U/L | 200 | 60.54±69.77 | 169 | 61.81±73.49 | 31 | 53.62±44.52 | 0.174 | 0.549 |
| AST, U/L | 207 | 30.26±20.1 | 174 | 30.61±20.56 | 33 | 28.41±17.62 | 0.843 | 0.565 |
| BUN, mmol/L | 194 | 6.11±5.66 | 155 | 4.77±1.94 | 39 | 11.43±10.55 | 0.000 | 0.000 |
| Cr, umol/L | 195 | 99.84±145.31 | 157 | 65.78±15.12 | 38 | 240.55±290.66 | 0.000 | 0.001 |
| UA, umol/L | 184 | 253±96.15 | 148 | 254.84±81.14 | 36 | 245.44±143.57 | 0.000 | 0.707 |
| Cys C, mg/L | 152 | 1.13±1.31 | 120 | 0.81±0.26 | 32 | 2.33±2.5 | 0.000 | 0.002 |
| K, mmol/L | 209 | 4.32±0.76 | 164 | 4.28±0.61 | 45 | 4.46±1.15 | 0.000 | 0.306 |
| Na, mmol/L | 208 | 139.4±4.27 | 164 | 140.1±3.61 | 44 | 136.81±5.47 | 0.000 | 0.000 |
| Cl, mmol/L | 208 | 102.42±3.78 | 164 | 102.61±3.75 | 44 | 101.72±3.83 | 0.747 | 0.167 |
| Ca, mmol/L | 208 | 2.07±0.22 | 164 | 2.09±0.22 | 44 | 1.98±0.21 | 0.959 | 0.003 |
| CK, U/L | 114 | 107.28±109.2 | 88 | 85.43±60.34 | 26 | 181.21±184.01 | 0.000 | 0.015 |
| CK-MB, U/L | 114 | 16.76±11.75 | 88 | 15.96±12.76 | 26 | 19.47±6.89 | 0.364 | 0.182 |
| Mb, ug/L | 71 | 53.31±105.29 | 57 | 29.44±29.19 | 14 | 150.51±208.21 | 0.000 | 0.049 |
| cTnI, ug/L | 52 | 0.02±0.02 | 43 | 0.01±0.01 | 9 | 0.04±0.04 | 0.000 | 0.061 |
| LDH, U/L | 49 | 222±71.78 | 37 | 203.11±70.31 | 12 | 280.25±37.42 | 0.085 | 0.001 |
| FBG, mmol/L | 58 | 7.95±4.17 | 50 | 7.3±3.62 | 8 | 12.05±5.27 | 0.368 | 0.002 |
| RBG, mmol/L | 45 | 8.36±4.51 | 40 | 7.95±3.91 | 5 | 11.57±7.75 | 0.064 | 0.091 |
| PT, s | 57 | 12.35±1.94 | 48 | 12.22±1.56 | 9 | 13.08±3.38 | 0.153 | 0.225 |
| APTT, s | 56 | 31.52±6.69 | 48 | 31.14±4.89 | 8 | 33.75±13.58 | 0.000 | 0.299 |
| INR | 57 | 1.17±0.18 | 48 | 1.16±0.15 | 9 | 1.24±0.32 | 0.160 | 0.222 |
| FIB, g/L | 57 | 5.08±1.46 | 48 | 4.94±1.47 | 9 | 5.81±1.21 | 0.938 | 0.102 |
| D-D, ug/L | 28 | 183.96±233.46 | 21 | 108.9±86.48 | 7 | 409.14±376.11 | 0.007 | 0.080 |

\*: Levene test was used for the homogeneity of variance test. Direct Bilirubin (DBil), Indirect Bilirubin (IBil), Total Bilirubin (TBil), Total Protein (TP), Albumin (ALB), Globulin (GLO), albumin-globulin ratio (A/G), Alanine Amiotransferase (ALT), Aspartate Aminotransferase (AST), Blood Urea Nitrogen (BUN), Creatinine (Cr), Uric Acid (UA), Cystatin C (Cys C), Kalium (K), Sodium (Na), Chloride (Cl), Calcium(Ca), Creatine Kinase (CK), Creatine Kinase-MB (CK-MB), Myoglobin(Mb), Cardiac Troponin I (cTnI), Lactate Dehydrogenase (LDH), Fasting Blood Glucose (FBG), Random Blood Glucose(RBG), Prothrombin Time(PT), Activated Partial Thromboplastin Time (APTT), International Normalized Ratio (INR), Fibrinogen (FIB), D-Dimer (D-D).

The ROC curve was used to analyze the hematological parameters with significant differences between the two groups. The parameters with AUC <0.6 and no statistical significance (*P*> 0.05) with AUC = 0.5 were excluded. Next, we analyzed the diagnostic efficacy of other hematology parameters (Table 4) in distinguishing moderate from severe COVID-19 cases. Taking the Youden index and the purpose of clinical screening, the best diagnostic cutoff was selected.

**Table 4.** ROC analysis for several significantly parameters.

| Parameters | AUC | SE<sup>a</sup> | P Value<sup>b</sup> | 95% CI of AUC |  |
| --- | --- | --- | --- | --- | --- |
|  |  |  |  | LL | UL |
| WBC | 0.652 | 0.054 | 0.009 | 0.546 | 0.758 |
| Neu# | 0.726 | 0.047 | 0.000 | 0.634 | 0.819 |
| Baso# | 0.622 | 0.059 | 0.037 | 0.506 | 0.738 |
| **NLR** | **0.890** | **0.033** | **0.000** | **0.824** | **0.955** |
| **PLR** | **0.842** | **0.040** | **0.000** | **0.763** | **0.921** |
| MCH | 0.636 | 0.048 | 0.020 | 0.542 | 0.730 |
| RDW_CV | 0.652 | 0.066 | 0.009 | 0.522 | 0.783 |
| RDW_SD | 0.757 | 0.056 | 0.000 | 0.648 | 0.866 |
| **NLR&RDW-SD** | **0.938** | **0.018** | **0.000** | **0.902** | **0.973** |
| **NLR&RDW-CV** | **0.923** | **0.022** | **0.000** | **0.880** | **0.967** |

a. Under nonparametric assumptions; b. Zero hypothesis: real area = 0.5. Area under Curve (AUC), Standard Error (SE), Confidence interval (CI), Lower Limit (LL), Upper Limit (UL);
$NLR\&RDW\_SD = 0.078737 \times NLR + 0.489253 \times RDW\_SD - 19.9587$
$NLR\&RDW\_CV = 0.1031 \times NLR + 1.1137 \times RDW\_CV - 14.5006$

The results showed that NLR was the best single parameter in distinguishing moderate and severe cases. For NLR, the AUC, the best cut-off value, the sensitivity and the specificity were 0.890, 13.39, 83.3% and 82.4% respectively when taking the Youden index and the purpose of clinical screening. Followed by PLR parameter, its AUC and best cut-off value, sensitivity and specificity of identifying moderate and severe COVID-19 patients were 0.842 267.03, 83.3% and 74.0% respectively. The combined parameters fitted by the LDA method were also used for the diagnostic efficacy analysis in the differentiation between the severe and the moderate groups. Among the parameters said above, it is found that the combined parameter NLR&RDW-SD generated by linear fitting of NLR and RDW-SD according to the formula below the table 4 had the best diagnostic efficiency (AUC is 0.938). When cut-off value was 1.046, the sensitivity of distinguishing severe type from moderate cases of COVID-19 was 90.0% and the specificity 84.7%. The second most effective parameter was fitting parameter NLR&RDW-CV (AUC = 0.923). When the cut-off value was 0.62, the sensitivity of distinguishing severe type from moderate cases of COVID-19 was 90.0% and the specificity 82.4%. When the combined parameter NLR&RDW-SD≥1.046 or NLR&RDW-CV≥0.62, it is more likely that the patient is severe type. The combined parameters are better than the single parameter, which can better assist the clinician to judge the patient’s condition.

Meanwhile, we used the cut-off values of the best single-parameter NLR and the combined parameter NLR&RDW-SD as the judgment thresholds and listed the fourfold tables (Table 5 and Table 6) to calculate the other diagnostic items, it was found that the combined parameter NLR&RDW-SD had better diagnostic performance, with a diagnostic accuracy rate of 85.7%, and better predictive value and likelihood ratio than the single parameter NLR.

**Table 5.** Fourfold table for differential severe cases with COVID-19(NLR)

| NLR |  | Gold Standard(Clinical confirmed) |  | Total |
| --- | --- | --- | --- | --- |
|  |  | Positive | Negative |  |
| NLR | Positive | 25 | 23 | 48 |
|  | Negative | 5 | 108 | 113 |
| Total |  | 30 | 131 | 161 |

**Table 6.** Fourfold table for differential severe cases with COVID-19(NLR&RDW-SD)

| NLR&RDW-SD |  | Gold Standard(Clinical confirmed) |  | Total |
| --- | --- | --- | --- | --- |
|  |  | Positive | Negative |  |
| NLR&RDW-SD | Positive | 27 | 20 | 47 |
|  | Negative | 3 | 111 | 114 |
| Total |  | 30 | 131 | 161 |

**Table 7.**
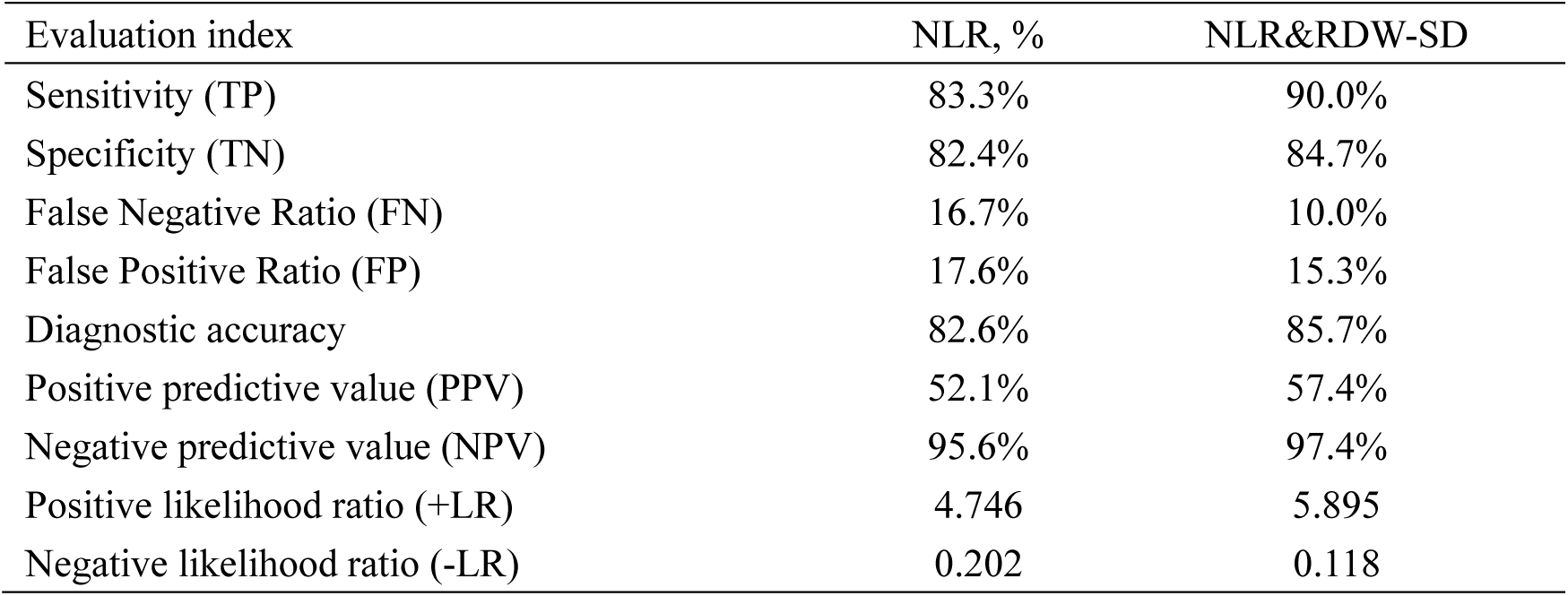
Diagnostic evaluation items of the best single and combined parameters.

## Discussion

The novel coronavirus (SARS-CoV-2) belongs to the beta-type RNA coronavirus. Like SARS-CoV that caused the outbreak of SARS in 2003 and MERS-CoV that caused the outbreak of MERS in 2012, it is different from the four human coronaviruses that previously caused the common cold in humans, and can cause severe respiratory diseases in humans [17]. Although the current epidemiological situation shows that the SARS-CoV-2 is more contagious than SARS-CoV and MERS-CoV, clustering of outbreak among people, but its lethality is less severe [35-39]. Compared with the outbreak of SARS in 2003 which caused 8098 confirmed diagnoses and 774 deaths (mortality rate 9.6%) in 37 countries [20] and the outbreak of MERS in 2012 which caused 2494 confirmed diagnoses and 858 deaths (mortality rate 34.4%) in 27 countries [21], COVID-19 outbreak also had a high mortality rate in the early stage in some areas. The first 41 confirmed infection cases admitted to hospitals in Wuhan from December 16, 2019 to January 2, 2020 showed that the COVID-19 patients had a 15% mortality rate at the beginning of the outbreak, and 32% of patients required ICU monitoring and treatment [11]. As of the latest data published by the WHO on March 10, 2020, the mortality rates of COVID-19 patients in different countries and regions were 3.88% in China, 5.05% in Italy, 4.03% in the United States, 0.75% in South Korea, 3.81% in Iran, 2.73% in Spain, 2.14% in France, 1.75% in Japan, 0.93% in Britain, 0.18% in Germany, etc. The global mortality rate (including the Diamond Princess) is 2.66% [19]. Therefore, many experts remind us that SARS-CoV-2 may coexist with humans for a long time [35] and the potential infection risk needs special attention with the increase in the number of asymptomatic infections [18]. Marc Lipsitch, a professor of epidemiology at Harvard University, predicted that the SARS-CoV-2 would infect 40% -70% of people worldwide by 2020 [34]. Therefore, tracking and screening of suspected cases are very important because that not all COVID-19 patients have symptoms such as fever or dry cough in the early stage and it is possible for some patients to develop into severe or critical cases. The current epidemic situation in China appears controlled, but the global epidemic outside China is still in the early to middle stages of the outbreak [36]. Neither the detection capability of viral nucleic acid kits nor the popularity rate of imaging CT [32] can support large-scale screening of all populations, so if the most conventional peripheral hematology test methods have characteristic changes or prompts for infected patients, especially those with severe infections, they will be very helpful for clinicians to intervene early to reduce the mortality of patients and relieve the pressure of the epidemic.

This study reviewed the epidemiological, underlying diseases and signs, as well as the changes in blood routine, biochemical, and coagulation test results of 45 patients with SARS-CoV-2 infection in different disease severity in Jingzhou, Hubei province, China. There was no significant difference seen in the epidemiological finding between two groups (*P*>0.05). For the underlying diseases, there were 4 patients with hypertension, of which 3 (30%) were severe type. Of the 45 patients, 40 (89%) had fever and 27 (60%) had dry cough, 19 (42%) had fatigue, 15 (33%) had chills and 13 (28.9%) had myalgia. Fever and dry cough were still the most common symptoms in patients with COVID-19. Although there are not many cases reviewed, but the trend is similar to that reported in the literature [26, 33]. In the comparison of hematological parameters, WBC, Neu#, NLR, PLR, RDW-CV and RDW-SD in the severe group were significantly higher than that in the moderate group (*P* <0.05); meanwhile, Lym#, Eos#, HFC%, RBC, HGB and HCT in the severe group were significantly lower than that in the moderate group (*P* <0.05). Among them, WBC and Neu # are significantly higher in severe patients. It may be related to the persistent infection and prolonged hypoxia, leading to compensatory hyperplasia of the bone marrow to release more granulocytes and the results are consistent with the findings of Chen et al [27]. The significant lymphopenia in the severe group may be caused by the SARS-CoV-2 continuing to invade more lymphocytes, proliferate and cause the lymphocytes to die even to become depleted when they reach the spleen and other immune organs. The lymphopenia has been reported by many scholars [9, 11, 14, 16, 25-28]. The same phenomenon was also seen in SARS-CoV and MERS-CoV infections [29-30]. The trend of LYM # we found is consistent with the description mentioned in the Diagnosis and Treatment Guidelines (Trial Version 6) [2]. A study of 1099 COVID-19 patients by the team led by Zhong Nanshan showed that the proportion of lymphopenia reached 82.1% [27]. There was no significant difference in platelet count between the moderate and severe patients (*P*> 0.05). When the SARS broke out in 2003 in Guangzhou, China, it was first reported that the while blood cells count was normal or decreased (80.2%), the lymphocytes and eosinophils decreased, monocytes increased and thrombocytes decreased in some patients [22]. Similarly, a report from Hong Kong indicated that of 157 patients 153 (98%) had lymphopenia, 87 (55%) had thrombocytopenia, 77 (49%) had thrombocytosis, and 95 (61%) had hemoglobin decreased more than 20 g/L. The autopsy showed a decrease in lymphocytes in haemato-lymphoid organs, and multivariate analysis showed that old age and high concentration of LDH were independent predictors of poor prognosis. Lymphopenia and T lymphocyte subpopulation depletion may be related to the disease [23]. The results of our study are similar to the reported above [3, 26 and 27]. Lymphocytes and platelets are also important indicators for monitoring the peak viral load and immunopathological damage in the palliative treatment of lung disease with Abidol combined with lopinavir and ritonavir [24]. In this study, both the ratio of neutrophils to lymphocytes (NLR) and the ratio of platelets to lymphocytes (PLR) in the severe group showed significantly higher (*P* <0.05), and showed the best single parameter differential diagnostic efficacy (NLR AUC = 0.890, PLR AUC = 0.842). Liu et al [3] from Beijing Ditan Hospital Capital Medical University (Beijing, China) also suggested that NLR was helpful for early detection of severe COVID-19 patients nad had a high prediction accuracy (AUC = 0.849), which is consistent with the conclusion of this article. In addition, this study found that the red blood cell parameters (RBC, HGB, HCT) were significantly reduced in the severe group, while the morphological parameters (RDW-CV, RDW-SD) were significantly higher in the severe group than moderate patients, which may result from the immune damage that leads to the suppression of the bone marrow, leading to the gradual increase of anemia that causes the compensatory hyperplasia of erythroid cell line, a large number of immature red blood cells released to the peripheral blood, the activation of red blood cell apoptosis and peripheral phagocytosis, therefore result in the increase of the red blood cell distribution width[31]. In the comparison of the biochemical and coagulation indexes tested, it was found that as the disease progressed, DBil, GLO, BUN, Cr, Cys C, CK, Mb, LDH and FBG in the severe group were significantly higher than that in the moderate group (*P* <0.05); meanwhile, ALB, Na ^+^ and Ca^2+^ in the severe group were significantly lower than that in the moderate group (*P* <0.05), which is also consistent with recent reports by most scholars [11, 16, 26-28].

According to the changing trend of NLR and RDW-SD, we use LDA to carry out linearly fitting on the different hematological parameters because when the development or diagnosis of a disease is affected by two mutually masked parameters, the diagnostic performance will be lower if only one parameter is used, for example one parameter with high sensitivity while the other with high specificity. The fitting of the two different parameters by ax + by + c = w into one combined parameter has the advantages of the two parameters in specificity and sensitivity at the same time and can be more effectively used for the diagnosis and prediction of diseases. In this study, after fitting analysis of different parameters, NLR&RDW-SD combined parameter is selected as the best indicator to distinguish moderate COVID-19 patients from severe cases. The AUC is up to 0.938 and the diagnostic accuracy rate up to 85.7%. The combined parameter can help clinicians prejudge the staging of patients and take effective treatment measures in advance.

This study is a single-center exploratory retrospective study. The included cases are only 45, of which 35 and 10 are moderate and severe patients respectively. Fever and dry cough are the most common symptoms. The age distribution is mainly in the 16-62 years old. Pregnant women, children and the elders, and asymptomatic patients were not included in the study.. Therefore, the specific application needs to be verified and confirmed by more clinical cases. At the same time, we did not analyze and study the prognosis of the included cases. However, this study suggests that it may be possible to find potentially severe patients through the most routine and basic hematological tests in the early stages of COVID-19 in order to provide patients with early clinical intervention, to reduce patient mortality and to help to control and prevent the epidemic. We believe that this study can provide some reference value for research and epidemic prevention in other countries and regions

## Summary

Currently, SARS-CoV-2 virus has begun to spread globally, but there is no clinically effective drug for COVID-19. Before SARS-CoV-2 vaccine can pass clinical trials and be widely and safely applied [35], it is inevitable that more patients will develop into severe or even critical patients. To establish an effective treatment strategy for severe and critical patients depends on early diagnosis and early warning of disease progression which is the key to reducing the overall mortality of patients with COVID-19 [25]. This study found that the combined parameter of NLR&RDW-SD can be used as an indicator to distinguish moderate COVID-19 patients from severe cases. The AUC is up to 0.938, based on its optimal cut-off value (1.046), the diagnostic accuracy is up to 85.7%, and there is a good positive and negative likelihood ratio. That is to say, if the result of NLR&RDW-SD of a COVID-19 patient exceeds 1.046, it suggests that there is a greater possibility that the patient’s situation is more likely to get worse or the patient is more likely to be a severe patient. If the result of NLR&RDW-SD is less than 1.046, it suggests that the patient is more likely get better or to be a moderate patient. This information will help clinicians to predict the severity and disease classification of patients, to take effective treatment measures in advance, to carry out differential treatment and to control the epidemic effectively.

## Data Availability

I declare that all data are true

## Acknowledgement

We thank all patients who participated in this study. Thanks to all health-care workers in our hospital for their efforts in caring for these patients. Thanks to all people who work so hard to fight against the novel coronavirus pneumonia (COVID-19).

## Statement of Ethics

This study was approved by Ethics Committee of Jingzhou Central Hospital. As this study was a retrospective study, there was no patient’s privacy data such as patient name, ID number, telephone and address were involved. Only demographic information and laboratory testing data of patients were collected and analyzed, so there is no informed consent in this study.

## Author Contributions

Changzheng Wang and Rongrong Deng have access to all data in this study and are responsible for the integrity of the data and the accuracy of the data analysis. Liyao Gou and Feng Shao are responsible for the research content, experimental design and the ethical approval, Zhongxiao Fu, Weiyang Fu and Xiao Ding for the data collection and data accuracy, Jianping Xiao and Guanzhen Wang for the statistical analysis, Tao Li and Xiaomei Zhang for the manuscript draft, and Chengbin Li and Xiulin Xiao for critical revision of the manuscript. All authors have reviewed and approved the final version.

